# Successful Application of Wastewater-Based Epidemiology in Prediction and Monitoring of the Second Wave of COVID-19 in India with Fragmented Sewerage Systems- A Case Study of Jaipur (India)

**DOI:** 10.1101/2021.09.11.21263417

**Authors:** Sudipti Arora, Aditi Nag, Aakanksha Kalra, Vikky Sinha, Ekta Meena, Samvida Saxena, Devanshi Sutaria, Manpreet Kaur, Tamanna Pamnani, Komal Sharma, Sonika Saxena, Sandeep K Shrivastava, A. B. Gupta, Xuan Li, Guangming Jiang

## Abstract

The present study tracked the city-wide dynamics of severe acute respiratory syndrome-corona virus 2 (SARS-CoV-2) RNA in the wastewater from nine different wastewater treatment plants (WWTPs) in Jaipur during second wave of COVID-19 out-break in India. A total of 164 samples were collected weekly between February 19^th^ and June 8^th^, 2021. SARS-CoV-2 was detected in 47.2% (52/110) influent samples and 37% (20/54) effluent samples. The increasing percentage of positive influent samples correlated with the city’s increasing active clinical cases during the second wave of COVID-19 in Jaipur. Furthermore, WBE based evidence clearly showed early detection of about 20 days (9/9 samples reported positive on April 20^th^, 2021) prior to the maximum cases & maximum deaths reported in the city on May 8^th^, 2021. The present study further observed the presence of SARS-CoV-2 RNA in treated effluents at the time window of maximum active cases in the city even after tertiary disinfection treatments of UV & Chlorine. The average genome concentration in the effluents and removal efficacy of six commonly used treatments; Activated Sludge Treatment + Chlorine disinfection (ASP + Cl_2)_, Moving Bed Biofilm Reactor (MBBR) with Ultraviolet radiations disinfection (MBBR + UV), MBBR + Chlorine (Cl_2_), Sequencing Batch Reactor (SBR) and SBR + Cl_2_ were compared with removal efficacy of SBR + Cl_2_ (81.2%)> MBBR + UV (68.8%) > SBR (57.1%) > ASP (50%) > MBBR + Cl_2_(36.4%). The study observed the trends & prevalence of four genes (E, RdRp, N, and ORF1ab gene) based on two different kits and found that prevalence of N> ORF1ab >RdRp> E gene, suggested that the effective genome concentration should be calculated based on the presence/absence of multiple genes. Hence, it is imperative to say that using a combination of different detection genes (E, N, RdRp & ORF1ab genes) reduce false positives in WBE.

**Graphical Abstract:** 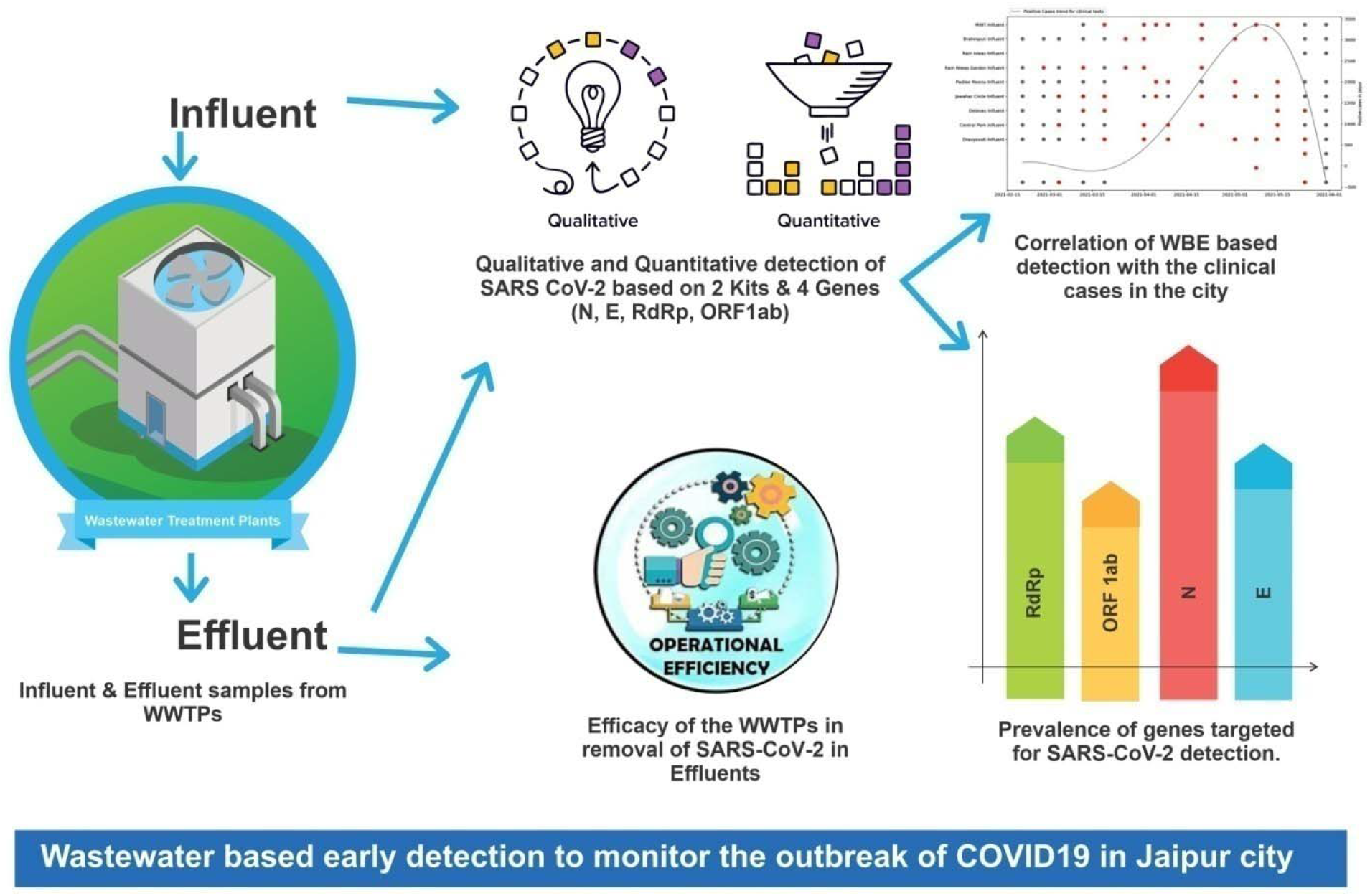

**Highlights:**

- Successful application of WBE with prediction of 14-20 days for COVID-19 in Jaipur
- A comparison of SARS-CoV-2 RNA removal efficacy of 9 WWTPs was investigated
- SBR showed better performance than MBBR with SARS-CoV-2 RNA removal from wastewater
- Presence of SARS-CoV-2 in effluents even after UV and Chlorine disinfection
- Using a combination of different detection genes reduce false positives in WBE

## Introduction

An outbreak of pneumonia of unknown etiology was first reported in Wuhan (Hubei province, China) in late 2019, and the metagenomics sequencing shed light on the association of this outbreak with a novel coronavirus (nCoV) (Mehta et al., 2020). The “novel coronavirus-infected pneumonia” was officially designated as COVID-19 caused by SARS-CoV-2 (Zhu et al., 2020; Gorbalenya et al., 2020). A total of 222,895,613 confirmed cases, including 4,602,961 deaths, were officially announced all over the world, by September 8^th^, 2021 (https://www.worldometers.info/coronavirus/), with distressing consequences on human health and economy, particularly in the United States, India, and Russia, among others (Johns Hopkins University and Medicine, 2021). The available epidemiological evidence strongly suggests that COVID-19 is primarily transmitted through respiratory droplets and contact routes (Rothan & Byrareddy, 2020). The tracing of SARS-CoV-2 genetic material— viral RNA—in stool and urine of COVID-19 patients (Chen et al., 2020; Peng et al., 2020; Young et al., 2020) shed light on the pattern of spread of virus dissemination by aqueous matrices. The circulation of virus was speculated to have occurred from malfunctioning sewage works (sewer networks and wastewater treatment plants (WWTP)) in the community (Zaneti et al., 2020; Ahmed et al., 2020).

Recently, there grew a huge interest in the scientific community in shedding of virus into feces as well as the presence and persistence of SARS-CoV-2 in health care and municipal effluents, although the potential of sewage to spread COVID-19 is extremely low and has not been reported to date (Ahmed et al, 2020; Medema et al., 2020). Both the World Health Organization (WHO) and the US Centers for Disease Control and Prevention (CDC) don’t consider COVID-19 as waterborne and finding clues to support this claim throughout literature has not reached a clear conclusion (CDC, 2020). The presence of SARS-CoV-2 in wastewater indeed raises the potential for sewage analysis to inform epidemiological monitoring of COVID-19 as wastewater-based epidemiology (WBE). WBE is regarded as a complementary approach for current clinical surveillance which includes providing information on the prevalence and spread of disease in a population (Bivins et al, 2020a, b). WBE based on raw wastewater fingerprinting to obtain qualitative and quantitative data within a given wastewater catchment, not only provides an early warning sign for disease outbreaks but also acts as a smart way of imposing preemptive quarantine (Sims et al., 2020).

For the last 1.5 years, several groups of researchers have been conducting different studies into sewage monitoring of SARS-CoV-2 with primarily two objectives. One is to detect the presence/surveillance of virus in a population for early epidemic prediction (SWEEP); and secondly, to assess infection risk to the public and sewage workers/operators from untreated/partially treated contaminated sewage and effluent as well as air surrounding wastewater treatment facilities (Tiwari et al., 2021). WBE is a potential tool to complement the current clinical surveillance as an affordable, convenient, and practical program as it gives a time period of at least 7-28 days in advance for early preparedness by providing information on the prevalence and spread of disease in a population which helps decision & policy makers for proper allocation of resources (Sims et al., 2021).

Considerable efforts have been devoted to detecting SARS-CoV-2 in sewage in several countries, particularly in high-and upper-middle-income communities such as the Netherlands, Italy, Spain, etc. where the sewerage systems are properly connected (Medema et al., 2020; La Rosa et al 2020; Randazzo et al., 2020;). However, the imbalance between the numbers of studies in developed countries, and those on the broad spectrum of developing and resource-limited communities especially India clearly indicates that much work has yet to be accomplished. Over 80% of wastewater is not connected to proper sewage networks and is discharged without treatment in India (CPCB Report, 2020). The problem with India’s sewerage system is that it is fragmented, and poorly connected. There is still a large percentage of the population that is not connected to any sewage treatment plants and sewerage infrastructure. The coverage of the sewerage system in Jaipur is less than that of the drainage system as it covers only 60 percent of the Jaipur municipal corporation area and caters to about 80 percent of the population (NIUA Report, 2019).

There are very few case studies which have been reported from India, across the nation, including i.e., Uttarakhand and Rajasthan from Northern India (Arora et al., 2020a, 2021), Hyderabad (Hemalatha et al., 2021; Kopperi et al., 2021) and Chennai from Southern India (Chakraborti et al, 2021); and Gujarat (Kumar et al., 2020a, 2021a, 2021b, 2021c) and Maharashtra from Central India (Sharma et al., 2021), most of which have successfully demonstrated the usefulness of WBE but on a limited scale. The awareness of WBE has been increasing during the COVID-19 pandemic; however, WBE is still not an established practice in developing regions like India. There are some challenges for an effective WBE implementation in India including lack of awareness among the public health officials and government authorities, leaders of corporations and the public. The implementation of a nation-wide WBE program in countries like India with a dissimilar sanitary coverage is an extremely complicated issue. The use of distinct sanitation systems, such as centralized sewer systems and on-site sanitation systems—pit latrines, bucket latrines and septic tanks—impose a challenge for WBE implementation in low and middle-income countries. The Viral RNA detection in dysfunctional sewer systems needs to be further explored (Gwenzi et al., 2021; Street et al., 2020).Therefore, it becomes even more imperative to validate these research in such systems to prove WBE as an efficient monitoring tool for early prediction.

Considering these limitations and challenges due to the fragmented sewerage system and the huge gap between generated versus treated sewage, this study aims to delineate how efficient can WBE be for predicting the upcoming surge of COVID-19 in Jaipur and whether such systems become a barrier in successful application of WBE? To answer these pertinent questions, the present research study was planned from Indian perspectives to bridge the knowledge gap between researchers, scientific community and government officials and policy makers and to successfully implement WBE at a city scale that could possibly help in controlling the pandemic. Thus the objectives of the present study were to (1) evaluate the implementation of WBE for Jaipur city, for prediction of second wave of COVID-19, (2) to determine the efficacy of different treatment systems in nine WWTPs in removal of SARS-CoV-2 loads, (3) determine and validate the prevalence of different genes involved using the combination of two kits and (4) standardizing the methodology including sample collection, transportation, pre-processing, etc. in a cost effective manner to establish WBE as an overall economical approach. Further, the study will substantiate the potential of WBE for the city-wide surveillance in Jaipur city to incorporate WBE into the regular monitoring programs and policy framework to manage the future COVID-19 wave efficiently.

## 2. Experimental Methodology

### 2.1. Wastewater Sampling

Influent and effluent samples were collected from nine municipal wastewater treatment plants (WWTPs) located across the Jaipur city for the monitoring of the second wave of COVID-19. The influent samples have been analysed for the prediction of second wave while the analysis of effluent samples was done for evaluating the efficiency of the WWTPs for the removal of viral loads. This is a longitudinal study wherein the samples were taken between February 20^th^, 2021 and June 8^th^, 2021. All the samples were collected as one Litre grabs in sterile bottles and transported to the Environmental Biotechnology Laboratory at Dr. B. Lal Institute of Biotechnology, Jaipur for further investigation and analysis. Appropriate precautions including ambient temperatures were taken in consideration for sample collection. Concerned personnel wore standard personal protective equipment (PPE) during the entire sampling process. The collected samples were transported to the laboratory at ambient temperatures of the city during the collection months, as adopted by Arora et al., (2020, 2021). Figure 1 shows the geographical locations of the sampling sites used in the study.

**Figure 1:**
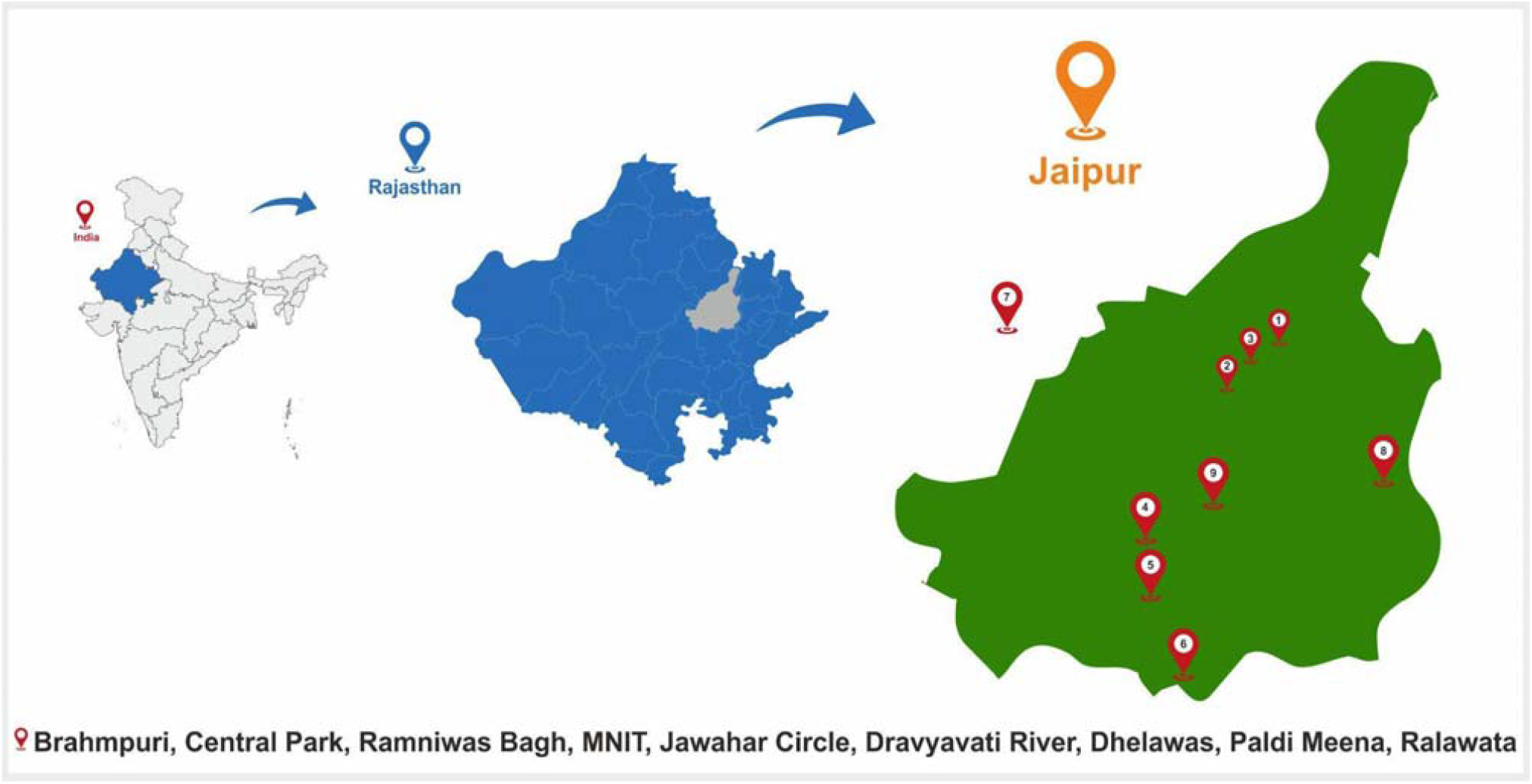
Sampling sites of Jaipur for the study.

### 2.2. Sample preparation

The samples for RNA isolation were prepared with slight modifications from the protocol described previously (Arora et al., 2020, 2021). The wastewater samples were surface sterilized using UV treatment for 30 minutes followed by manual mixing. Further 1 ml sample was aliquoted and centrifuged at 7,000 rpm for 30 minutes (for removal of debris & unwanted materials) and the supernatant was then processed for RNA extraction as described in Arora et al., (2021).

### 2.3. Viral RNA Extraction

Viral RNA was extracted from the processed wastewater samples via the MagMAX Viral/Pathogen Nucleic Acid Isolation Kit (Applied Biosystems) following the manufacturer’s instructions using the automated KingFisher™ Flex machine. The protocol involves “three wash” steps for the extraction of the RNA. Samples were vortexed for 10 seconds and then mixed with the extraction buffer consisting of binding solution, binding beads and Proteinase K (referred to as extraction master mix), vortexed for 30 seconds and then processed using the automated system. This is followed by three washing steps using Wash Plate 1 consisting of wash buffer, wash plate 2 and 3 each consisting of varying amounts of 80% PCR grade ethanol. The RNA is finally eluted out and the process takes about 24 minutes each time. The eluted RNA in the plates are then sealed and stored at -20°C till further use.

### 2.4. Qualitative and Quantitative detection of SARS-CoV-2

The qualitative and quantitative presence of SARS-CoV-2 RNA was detected in the total RNA extracted from the wastewater samples using CFX 96 Thermal Cycler (Bio-Rad) RT-PCR machine using two separate commercially available kits. Kit 1 was Allplex™ 2019-nCoV Assay RT-PCR, used for qualitative detection of SARS-CoV 2 consisted of 2019-nCoV MOM (prepared master mix), 5X Real-time One-step Buffer, Real-time One-step Enzyme and exogenous Internal Control (IC). The kit 1 targeted E gene, N gene and RdRp gene with FAM and HEX as internal controls to be read on Cal Red 610 and Quasar 670 fluorophore channels, respectively. The PCR reaction was set up by mixing 11 μL of the isolated RNA with 14 μL of RT-PCR master mix. The reaction protocol consisted of 1 cycle at 50°C for 20 min, 1 cycle at 95°C for 15 min followed by 45 cycles of denaturation at 94°C for 15 sec and combined annealing and extension for 30 sec at 58°C followed by plate read and detection. The PCR run was analyzed with Bio-Rad CFX Manager software version 3.1 (Bio-Rad Laboratories). As per manufacturer’s instructions, the detection of a minimum of any two genes (out of three) in a sample was considered positive based on Ct values.

To further quantify the presence of SARS-CoV-2 viral genome in the wastewater samples, InnoDetect One Step COVID-19 (Kit 2) was used wherein two different plasmid DNA consisting of N gene and ORF1ab gene separately was used to prepare a standard curve (a range of 10pg/μL to 0.01fg/μL) as per the protocol in the manufacturer’s instructions. These standard curves were then used for the quantification of the respective genes in the samples. RNase free water was used to make a main stock of concentration of 40ng/μL. The kit 2 consists of a master mix, primer probe (N gene, ORF1ab & RNaseP) and uses three fluorophore channels (HEX/VIC, FAM & ROX/Texas Red, respectively) for individual identification. Viral RNA of SARS-CoV-2 was used as a positive control and DNase RNase free water as a negative control provided with the kit. The reaction cycle consists of a reverse transcription step at 42°C for 15 min 1 cycle, cDNA initial denaturation at 95°C for 3 min 1 cycle, denaturation at 95°C for 15 Sec and combined annealing and extension at 60°C for 40 sec followed by plate read and detection. The samples with quantitative presence of any of the two genes (N or ORF1ab) or both the genes were considered positive.

### 2.5. Statistical Analysis

The co-detection of genes using different kits and the removal efficiency due to different treatment approach were visualized using R (ver. 3.31, http://www.R-project.org/). To evaluate the temporal effect, the viral concentration data (viral loads/positive detection rate) were paired with 7-day averaged new cases for Jaipur & India.

## 3. Results

### 3.1. Characteristics of selected sampling sites for prediction and monitoring of the second wave of COVID-19

Similar to other tier-2 cities of India, Jaipur also has a fragmented sewerage network system with different centralized and decentralized wastewater treatment plants (WWTPs). The main objective of the present study was to determine whether a WBE based early warning system can be established in such a fragmented system. Seven different sampling sites (Site 1, 2, 3, 4, 5, 6 and 8) were selected across the whole length and two sites (Site 7 and 9) were selected across the cross-section of the city, the details of which are described in Table 1. This ensured coverage of about 60-70% of the city population connected to the main sewerage trunk. Of the nine sites, one (Site 4) was a system connected to an academic institution (MNIT Jaipur) connected with a residential capacity of 2000 inhabitants. Site 2, 3, 5 and 8 are small-sized decentralized systems which receive wastewater from multiple catchment areas and inhabit population size of about 5000 individuals. Site 1 and 9 are medium-sized decentralized systems with a population size of greater than 50,000 individuals while site 6 and 7 are large centralized systems with population size of about 5 Lakhs (official data obtained from Jaipur Development Authority (JDA), & NIUA report 2021).

**Table 1:** Details of the sampling location sites along with treatment characteristics of WWTPs located in Jaipur, Rajasthan

| Site No. | Sampling Location | Type of Secondary treatment technology | Type of Tertiary Treatment | Dosage & contact time of tertiary treatment | Design Capacity (MLD) | Flow Rate (avg. MLD) | Number of connected residents (Approx.) |
| --- | --- | --- | --- | --- | --- | --- | --- |
| Site 1 | Brahmpuri, Jaipur<br>26.9373°N,<br>75.8250°E | SBR | No treatment | NA | 27 MLD | ~8 | ~59,000 |
| Site 2 | Central<br>Park<br>Garden,<br>Jaipur<br>26.9048°N,<br>75.8073°E | SBR | Cl<sub>2</sub> (Bleach<br>Powder) | 4 ppm by<br>dropping<br>system | 1 MLD | ~1 | □7,000 |
| Site 3 | Ramniwas<br>Garden,<br>Jaipur<br>26.8963°N,<br>75.8100°E | MBBR | UV | NA | 1 MLD | ~1 | □7,000 |
| Site 4 | MNIT,<br>Jaipur<br>26.8640°N,<br>75.8108°E | MBBR | Cl<sub>2</sub><br>(Hypochlori<br>te) | 2.5-3 ppm,<br>30 min | 1 MLD | ~1 | □2000 |
| Site 5 | Jawahar<br>Circle<br>Garden,<br>Jaipur<br>26°50'29"N<br>,75°48'0"E | MBBR | UV | NA | 1 MLD | ~1 | □7,000 |
| Site 6 | Dravyavati<br>River,<br>Jaipur<br>26.7980°N,<br>75.8039°E | SBR | Cl<sub>2</sub><br>(Hypochlori<br>te) | 3-5 ppm,<br>30 min | 65 MLD | ~65 | □480,000 |
| Site 7 | Dhelawas,<br>Jaipur<br>27.3735°N,<br>75.8926°E | ASP | No<br>treatment | 3 ppm,<br>30 min | 65 MLD | ~62.5 | □480,000 |
| Site 8 | Paldi<br>Meena,<br>Jaipur<br>26.8759°<br>N,<br>75.8945° E | SBR | No<br>treatment | NA | 3 MLD | 0.6 – 0.7 | ~5,000 |
| Site 9 | Ralawata,<br>Jaipur<br>26.76873°<br>N,<br>75.93092°<br>E | ASP | Cl<sub>2</sub><br>(Hypochlorite) | 10 kg per<br>hour | 30 MLD | 20-22 | ~1,70,370 |

Note: MNIT = Malaviya National Institute of Technology, MBBR= Moving Bed Biofilm Reactor, SBR= Sequencing Batch Reactor, ASP = Activated Sludge Process, Cl<sub>2</sub>= Chlorine disinfection, UV- Ultra violet disinfection, MLD= million litres per day, NA= Not applicable

Grab samples were collected every week during the entire duration of the study. Samples were collected from the sites located at the centre towards the sites at the upstream or downstream across the sewage trunk line. As a result, samples of the sites closer to the centre were collected around 11 AM while those of the sites at the terminal were collected around late afternoon at 1 PM. Wastewater sample collection from WWTPs, its transportation to the experimental laboratory and pre-processing before RNA extraction is a challenge in terms of both logistic feasibility as well as a for the applicability of WBE at a city scale. Our previous studies (Arora et al., 2021) have already reported that direct RNA extraction (without pre-processing) from 1 ml centrifuged supernatant of the properly mixed 1 Litre collected wastewater sample is sufficient enough for the qualitative detection of SARS-CoV-2. As a result, the similar protocol was used in this study.

### 3.2. Qualitative and Quantitative detection of SARS-CoV-2 in the Influent samples of WWTPs with monthly variations and correlation with the active cases in the city

In the present study, we reported weekly data of wastewater samples collected from nine different locations for sixteen weeks during February to June 2021 and the results are mapped in heat map as shown in Figure 2. The average Ct values for E, RdRp, N and ORF 1ab genes were 32.3, 35.1, 33.4 and 34.7, respectively. Likewise, the average Ct value of internal control (MS2 bacteriophage) was 27.3, and no SARS-CoV-2 genes were detected in the negative control samples. We detected and quantified monthly variations in SARS-CoV-2 RNA from wastewater samples to understand the pandemic situation during the second wave in Jaipur, Rajasthan (India). The longitudinal analysis of the wastewater samples collected from the nine sites showed the first detection of SARS-CoV-2 as early as 27^th^ February 2021 as evident from Figure 2. The percentage prevalence of positive influent samples to the total samples collected showed 44.4% positivity on 19^th^ March 2021 when the new active case number per day was only 61. The percentage positivity then increased to almost 100% from 26^th^ March 2021 and continued till 15^th^ May 2021 before declining. The increasing prevalence of percentage positive influent wastewater samples correlated well with the increasing cases & deaths reported during the second wave of COVID-19 in Jaipur. The noticeable increase in the case number *viz*. 528 per day appeared on 5^th^ April 2021 which is 2 weeks post the significant number of positive wastewater samples (on 26^th^ March 2021). So, this time period of 2 weeks could be sufficiently utilized to control the ever increasing cases & deaths, in the city. This can also be correlated with the number of deaths due to COVID-19 in this duration wherein the 7 day moving death average was around 1 on 1st April which increased to 55.57 on 8^th^ May 2021 (peak of COVID-19). The restricted movement was imposed in the city on 17^th^ April 2021 when the new active case number had already reached 1484 per day, which rose to a maximum of 4202 on 7^th^ May 2021 (as per official data from www.covid-19india.org).

**Figure 2:**
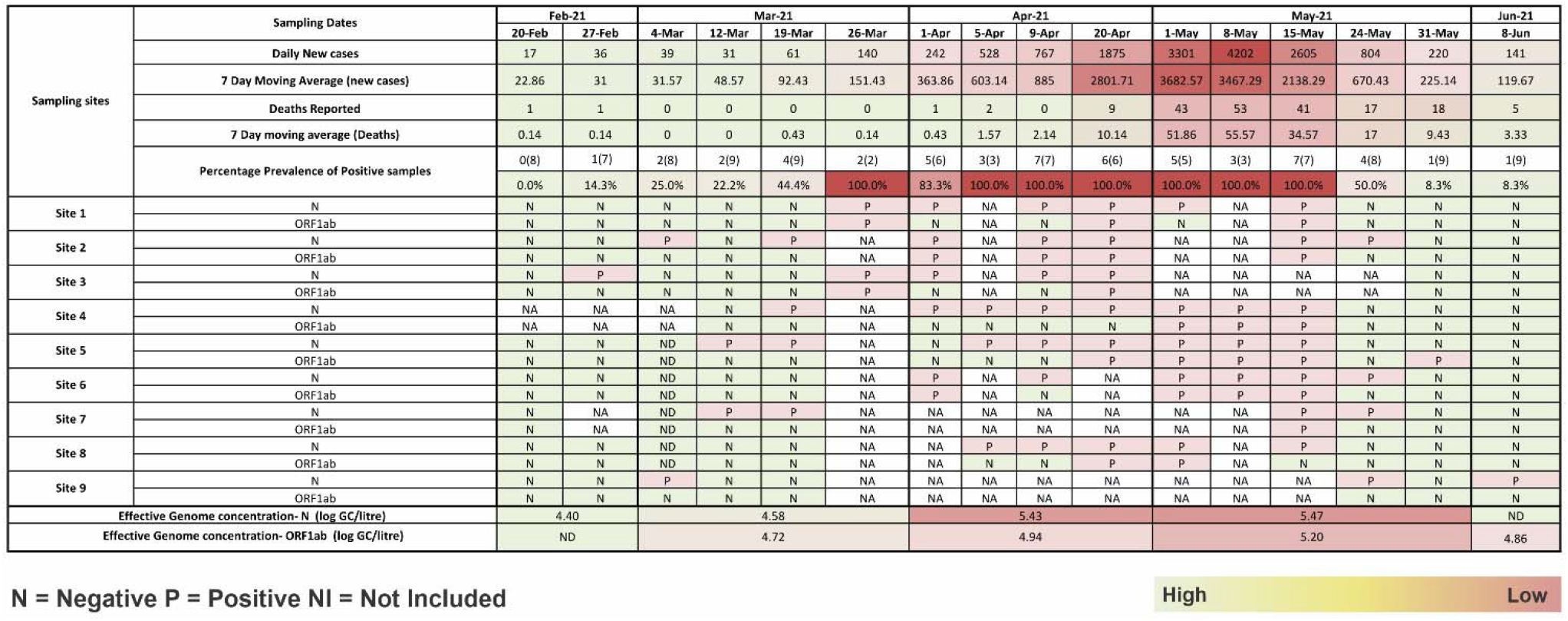
Heat map showing Temporal variation in positive prevalence of the SARS-CoV-2 targeted genes in Influent samples at various locations in Jaipur city with increasing active cases and deaths reported (Green N= negative, Pink P= Positive, NA= Not applicable)

The quantitative analysis of all the influent samples was also carried out wherein the genome copy number of N gene and ORF1ab gene was calculated. As observed in Figure 2, during the months of February, average genome concentration was log_10_ 4.40 GC/litre which increased to log_10_ 4.58 in March, to log_10_ 5.43 and 5.47 in April & May, respectively. The increasing genome concentration correlated well with the increasing number of active cases and mortality rate. Figure 3 also shows the detection of N gene as early as 27^th^ February 2021 coinciding with the qualitative analysis while ORF1ab gene was first detected on 26^th^ March 2021. Thus, the N gene could be detected 20 days prior to the significant rise in the new active patients per day while ORF1ab was detected 10 day prior. It is worth noting that the genome copies of both the genes were quantifiable in the wastewater samples throughout the second wave. Another interesting observation to note here is that although the N gene could be detected earlier during the rise of the cases, it was the ORF1ab gene which could be detected and quantified when the patient case numbers declined while the detection of N gene had already reached below limit of detection (LOD). The SARS-CoV2 RNA concentrations in the wastewater influent samples calculated from the N gene ranged from 4.4 to 6.04 log_10_ GC/L and ORF1ab ranged from 4.5-5.60 log_10_ GC/L (n = 110). Normalized viral loads of quantifiable wastewater influent samples from WWTPs were plotted and compared with new cases from Jaipur city and India (Figure 3).

**Figure 3:**
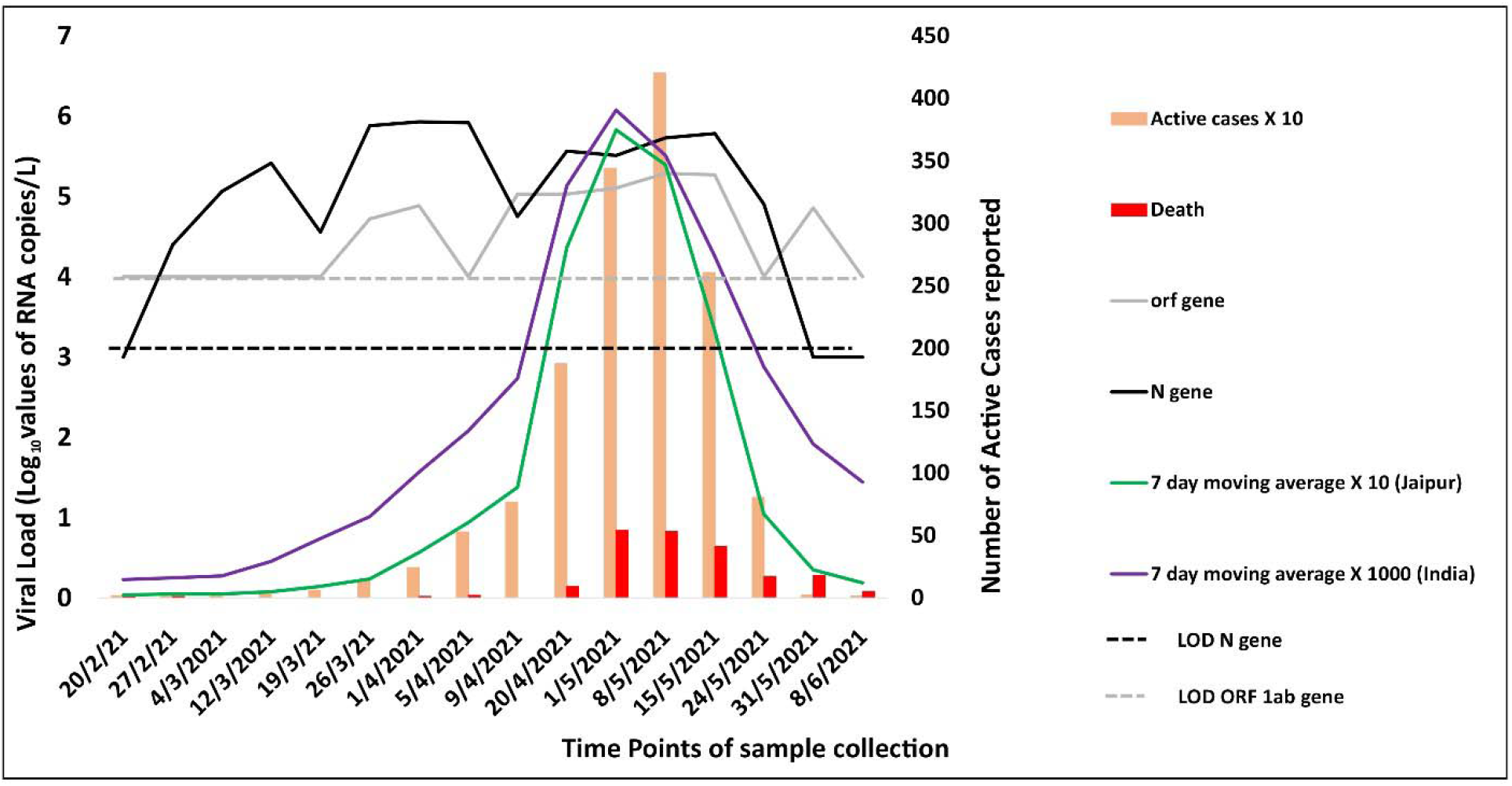
Analysis of SARS-CoV-2 concentration week wise with daily COVID-19 cases & Deaths reported in Jaipur & India (LOD-Limit of detection)

### 3.3. Prevalence of genes targeted for SARS-CoV-2 detection

Owing to the low sensitivity of the RT-PCR kits towards wastewater samples, two different kits consisting of five different probes were used for the study to ensure accurate detection. As already described in the methodology section, four different genes (N, RdRp, E and ORF1ab) were analysed where RdRp and E gene were detected qualitatively, ORF1ab was detected quantitatively and N gene was detected both qualitatively and quantitatively using two different probes. Figure 4 describes the prevalence & co-prevalence of four genes in both the samples (influent & effluent). Out of the 164 total wastewater samples tested, all the four target genes could be detected in only 15 wastewater samples whereas in other samples, genes were detected in different combinations. N gene was the most commonly detected gene in the samples wherein 33 samples tested positive for only one of the N gene targets (N detected by either or both probes from Kits 1 and 2) followed by 5, 8 and 12 samples in combination with E, RdRp and ORF1ab genes, respectively. Furthermore, one samples each consisting of positive targets of only ORF1ab or only RdRp genes. However, interestingly, the E gene showed the highest number of false positives and was never detected alone. Similarly, 16 and 7 samples were found positive for a combination of (N, RdRp, E gene) or of (ORF1ab, N and E genes), respectively.

**Figure 4:**
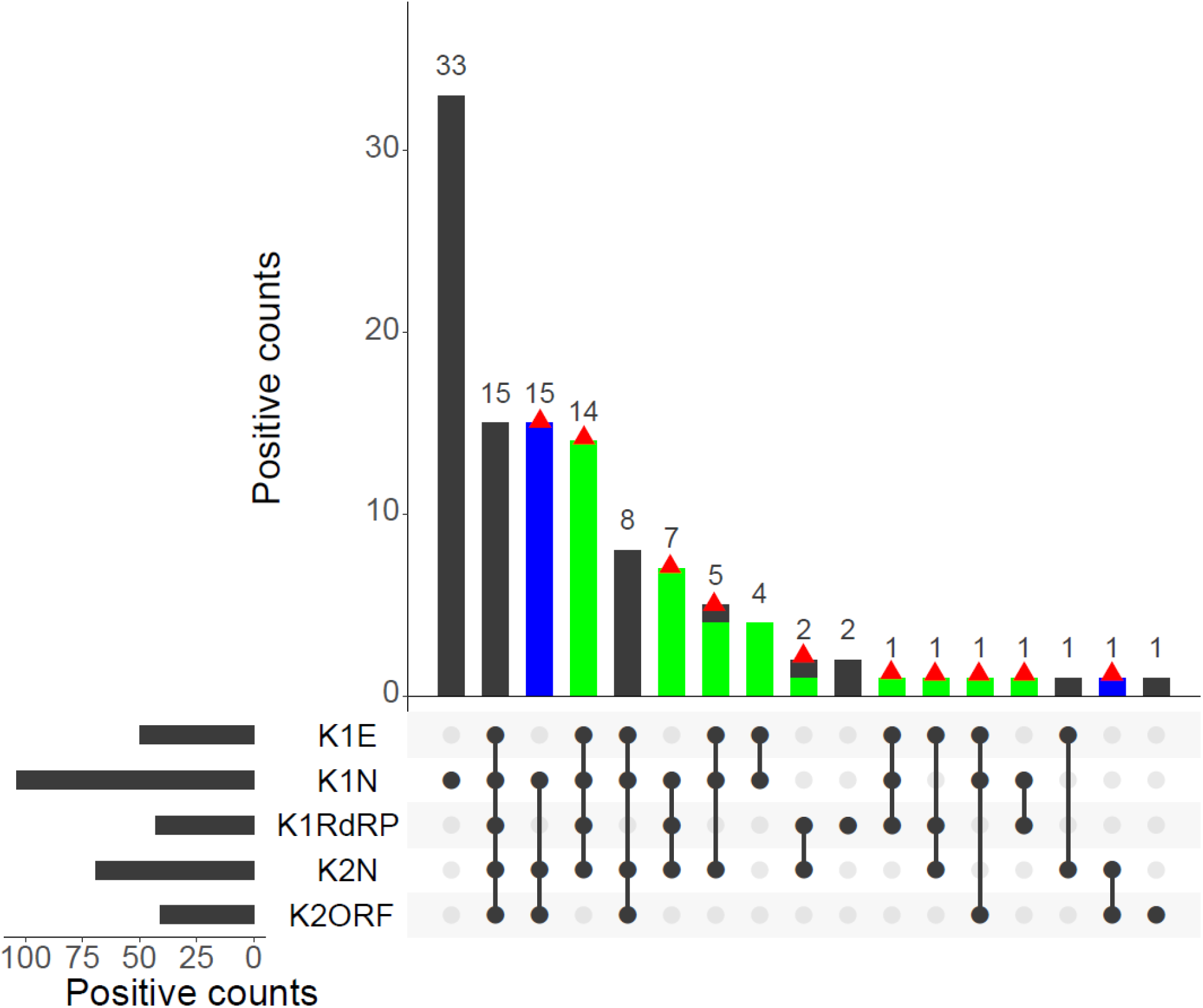
Positive detections using two kits with different genes (i.e., E, N, RdRp, ORF1ab) of all samples (both influent and effluent). The red triangle indicates positive samples identified based on the criteria. The blue and green bars indicate the false negative identified by Kit 1, and Kit 2, respectively.

Among the 164 wastewater samples analysed in the study, only 52 samples (30.5%) could be confirmed by both kits. However, the total number of wastewater samples that could be ruled positive by either of the kit target criteria was 72. The difference of 69.5% positive prevalence in samples was reported using combination of 2 kits. It is also observed that six samples were detected positive qualitatively, i.e., showed any two out of three genes positive using Kit 1 (including N gene) but could not be quantified by Kit 2. This could be attributed to the variation in the probe used for detection, sensitivity, and the detection limits of the two kits. In addition, 16 samples which were considered negative during the qualitative detection by Kit 1 (as per manufacturer’s criteria) were detected positive by Kit 2 (either N gene or ORF1ab gene or both present). These observations thus support the use of a combination of kits to achieve a finer distinction and broader detection of SARS-CoV-2 genome when compared to detection by a single kit. Hence, it is imperative to say that wastewater surveillance-based data must not be validated based on a single particular gene of SARS-CoV-2 but its effective gene concentration including multiple genes.

### 3.4. Efficacy of WWTPs in removal of SARS-CoV-2

The efficacy of the WWTPs in the removal of SARS-CoV-2 from the wastewater samples was observed in our earlier study during the first wave of COVID-19 (Arora et al., 2020, 2021). However, in the present study, it was observed that the efficacy of the WWTPs was compromised wherein the virus load could be detected in the effluent samples as well. Due to limitations (mainly permissions or breakdown of WWTP), in sample collection, the effluent samples from site 9 were not considered for analysis. Figure 5 summarizes the positive prevalence & the efficacy of different treatment technologies in removal of SARS-CoV-2 in the samples. It was observed that during March, an average of only 20% effluents samples were positive, which increased to 43.2% in April, and 55% in May. It was observed that this percentage can be correlated with the high active case-loads in the city, (between 9^th^ April 2021 and 24^th^ May 2021 as per www.COVID-19.org) which makes it difficult for WWTPS to remove SARS-CoV-2. The removal efficacy of different treatment technologies was also compared in terms of qualitative detection. Paired t-tests between the influent and effluent wastewater samples, taken on the same days, were performed to understand the significance of the SARS-CoV-2 gene removal efficacy of each treatment process, i.e., Activated sludge treatment ASP + Cl_2_, Moving Bed biofilm Reactor (MBBR) with Ultraviolet radiations (UV), MBBR + Chlorine (Cl_2_), Sequencing Batch Reactor (SBR) and SBR + Cl_2_ (Figure 5). Overall comparison of SARS-CoV-2 genome removal efficacy of different treatments is expressed on the total positive prevalence obtained throughout monitoring. The significance of SARS-CoV-2 genes removal efficacy in different treatment technologies includes the order of SBR + Cl_2_ (81.2%)> MBBR + UV (68.8%) > SBR (57.1%) > ASP (50%) > MBBR + Cl_2_ (36.4%).

**Figure 5:**
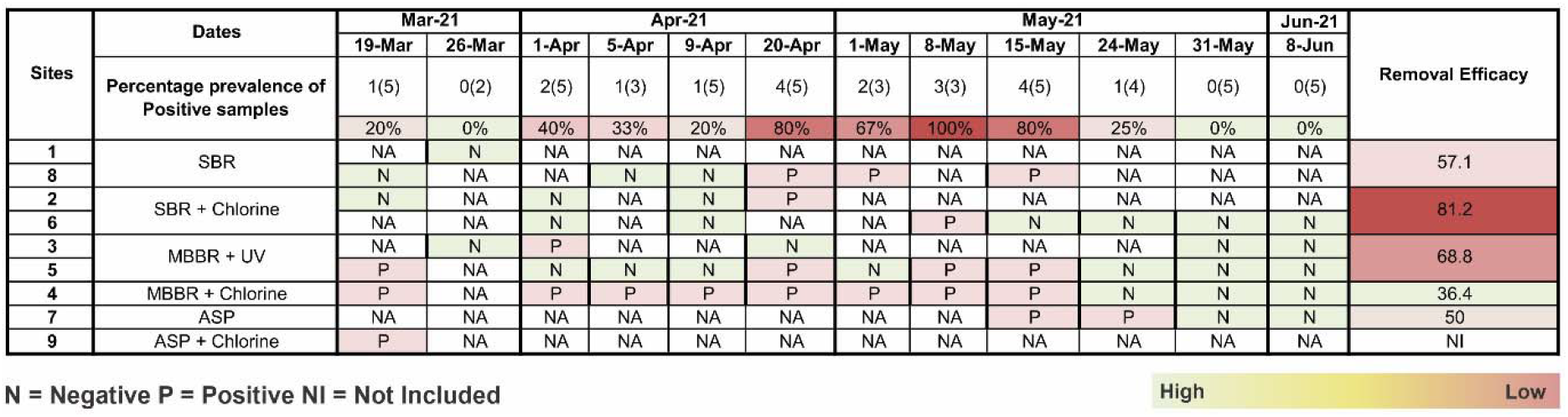
Temporal variation in gene copies of the SARS-CoV-2 targeted genes and effective gene concentration in Effluent samples at various locations in Jaipur city with removal efficacy of different treatments

The removal efficacy of the WWTPs was also analysed by comparing the viral genome load in terms of quantitative analysis of N gene and ORF1ab gene, respectively in the influent and the effluent samples. Figure 6 shows the box plots, of different treatment types using N gene & ORF1ab gene, respectively. These results suggested that in case of the sites like Sites 4 and 5 once the genome load in the influent samples exceeded the log of five, effluent loads were in the median range of 5 logs for both genes irrespective of the loads observed in corresponding effluent grabs which fluctuated between logs of 4 to 6.

**Figure 6:**
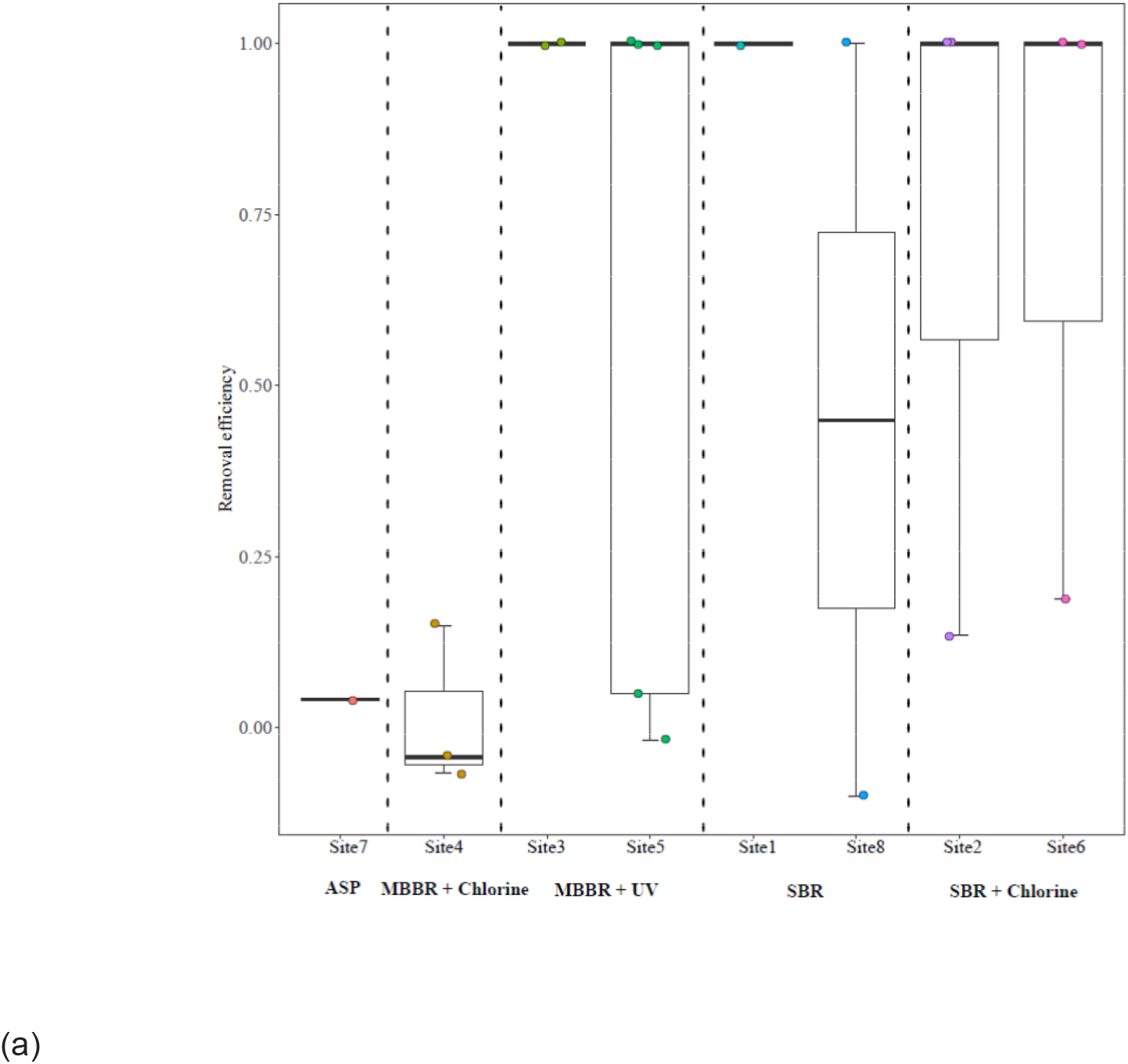
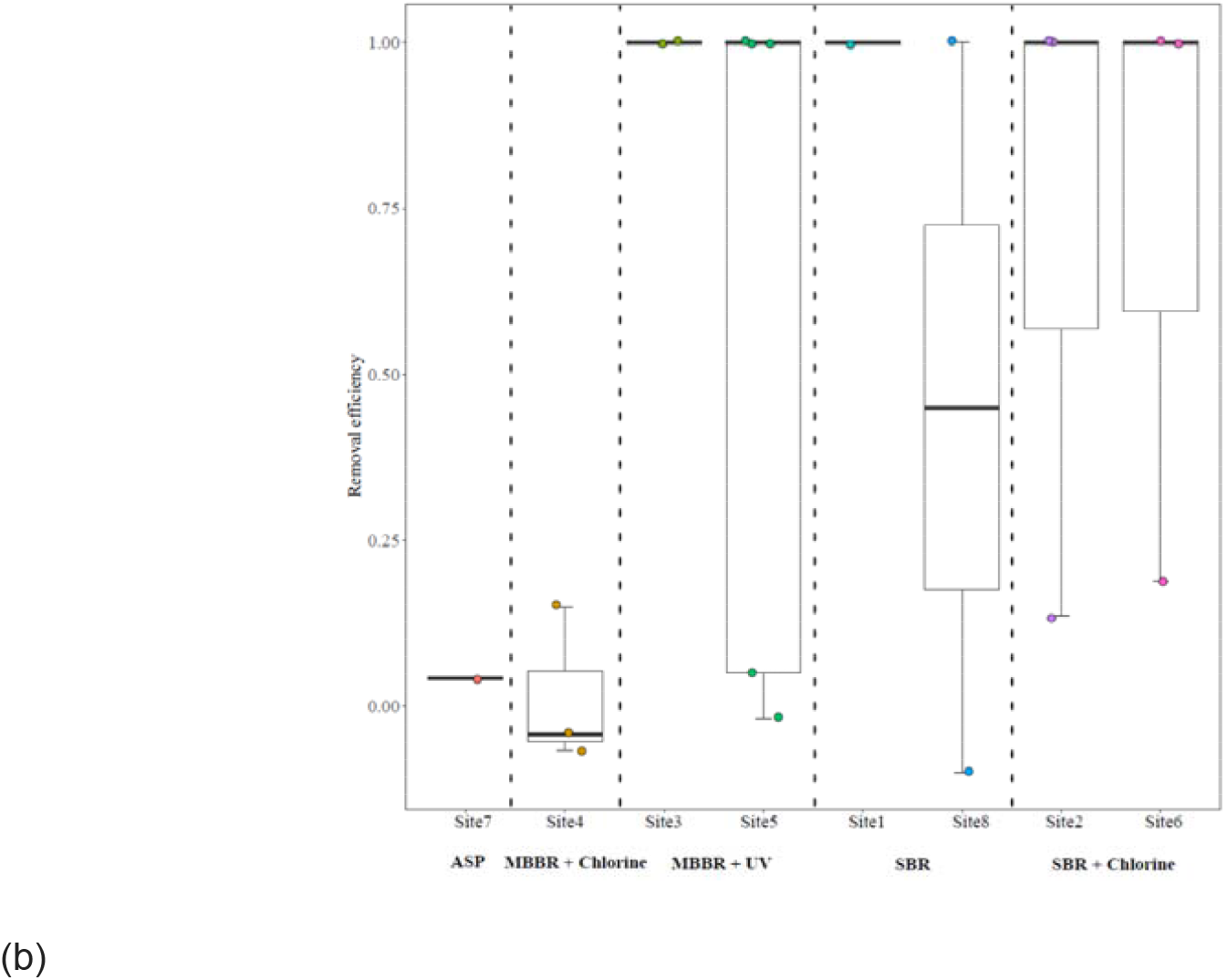
Box plot indicating Removal efficiency of SARS-CoV-2 RNA in wastewater in 5 different treatments systems at WWTPs using (a) N gene and (b) ORF1ab gene. Only the WWTP with no less than 2 data points for the removal efficiency was included in the figure.

## 4. Discussions

### 4.1. Sampling from fragmented and selected areas of a city can successfully be harnessed into a prediction model

WBE has been evaluated as a potential tool for the prediction of COVID-19 pandemic across various sections of the world (Fongaro et al., 2020; Ahmed et al., 2020). However, those studies have been carried out either based on a small scale in developing country (such as in studies of Kumar et al., 2020a, 2021a,b,c) or within a city with a highly connected sewer network in developed countries. The wastewater systems in majority of Indian urban cities are fragmented and composed of various decentralized treatment plants scattered throughout the city confines. The rural & slum areas and outskirts of these urban areas have practically a non-existent WWTP and thus most of the wastewater is dumped either in surface water bodies like rivers or in the community wide septic tanks. Given the fragmented state of the sanitation network, a mere ability to detect pathogens or biomarkers in wastewater is not sufficient. This detection needs to be carried out in a systematic, regular, and planned manner to monitor the community uniformly and warn the same well in advance about any potential threats. There is an urgent need for time-series data of SARS-CoV-2 RNA concentration in the wastewater that can be matched with the actual clinical survey data to confirm the utility and predictability of wastewater surveillance in India. This is also imperative for the adaptation of the Surveillance of Wastewater for Early Epidemic Prediction (SWEEP) on the policy level. This becomes more important in India since the development of a proper and integrated wastewater treatment system is far-fetched even in the urban areas considering the overall limitations. Therefore, the present study aimed to investigate the applicability of WBE in the prediction and monitoring of COVID-19 wave in a city level paradigm with a limited interconnected sewerage system. Despite the presence of disconnected and fragmented WWTPs, undergoing the treatment of only 60-70% of the total wastewater generated, the collection sites were selected such that they covered most of the total WWTPs installed in the city. A combination of small and medium decentralized WWTPs and large centralized treatment plants was selected to investigate in detail about the ability & feasibility of WBE to detect the upcoming COVID-19 active case load rise in advance. As mentioned in results section 3.1, it was observed that even with such a restricted coverage, the increase in positivity from various sites could be observed at least 14-20 days (at a total active case count of less than 50 per day) before a visible rise in newly detected active cases. Another important observation to be made in this case study that in contrast to Kumar et al., 2021 this study shows that if the sites are selected carefully it is possible to directly correlate the positivity rate of the sites to the upcoming wave in advance. As is the case in point where this study was able to predict the upcoming wave of COVID19 in Jaipur city, well in advance of 14-20 days (on 5^th^ April 2020) while the cases started increasing exponentially after 20^th^ April, with peak maximum cases load on 8^th^ May 2021. Overall, it can be highlighted that even in Indian sewage system networks rudimentary as they might be, WBE can be either directly (as observed here) or at least indirectly can be applied for early predictions (Kumar et al.,2021). Similar to the temporal variations in copy number reported in previous study by Kumar et al., 2021, an exact trend of copy number could not be established however, it was observed that the general positivity rates observed in this study could be correlated with the rise of caseloads in the city. Thus, it may seem that in a carefully monitored city, just qualitative detection might be enough in raising a rudimentary alarm for the city officials.

Another interesting observation was, in contrast to the previously published reports which suggest that WBE detection of SARS-CoV-2 genomes outlasts the clinical detection and falls slowly (Nemudryi et al., 2020); this study reported a sharp decline in positive prevalence rate (Figure 2 & 3) in the wastewater samples with the fall in cases. Within 2-3 weeks of COVID-19 second wave peak, the positive prevalence rate dropped to 50% (4/8) and 8% (1/9) in samples collected after 20^th^ May 2020. This contrast is interesting and needs to be further investigated. One of the possible explanations for this observation may be the limited sampling from selected collection points. Another explanation of this observation may be the symptoms present in the passing wave where coughing subsides contributing to decreased ratio of sputum to fecal viral load. However, more investigation is needed in this direction and in absence of a certain load to case number conversion metric, this observation might not be completely explained.

### 4.2. A combination of kits with multiple target probes has a large coverage and detection sensitivity

The premise of using WBE successfully in community wide surveillance of a disease or a biomarker is the shedding of respective pathogens or certain specific molecules into the wastewater (Weidhas et al., 2021). SARS-CoV-2 is known to be shed in wastewater through body excreta of any infected individual regardless of the manifested symptoms. Although several targets for detection of such diseases are available, several factors contribute to the efficiency of the method such as sensitivity of a detection primer-probe combination or any inhibiting factor present in each test sample. Therefore, it can be argued that relying on only one type of target or probe may lead to false negative or positive detection. Since timely detection of the target is essential for the management of the disease in the community, the percentage efficacy of two different kits was compared. To surveil and continuously monitor the presence of the viral particles, already available and ICMR approved kits were used as a part of investigating the uniform up-scalement of the city-wide surveillance. The set of targets consisted of both structural (N, E) and non-structural (RdRp and ORF1ab) genes (Naqvi et al., 2020). Out of total collected 164 samples, 148 samples were confirmed positive by kit 1 which could also be quantified by kit 2. However, 16 samples were confirmed positive only based on 2 kits used. Interestingly, only 30.48% (50 out of 164) of the positive samples could be detected by both the kits while 12.8% (21 out of 164) of the samples were additionally identified by either Kit 1 or Kit 2 alone.

It was also observed that most of the samples uniquely detected by Kit 1 were collected from a site connecting to centralized wastewater treatment plants (Sites 7 and 8) while all the samples which could only be detected by Kit 2 were from decentralized systems (Sites 1, 3 and 5). The sites which show sample variability between the detection by Kit 1 versus Kit 2 operate at a different scale and collect over different sizes of catchment area. Thus, the variability in detection could be because of the differential sensitivity of the primer probe set used in Kit 1 and 2. Or it could be due to the difference in silt or contaminant levels accumulated during the collection of wastewater in larger catchments between the origin and the centralized treatment plants. Furthermore, the sites which could be ruled positive only by Kit 2 were tested positive for at least one of the three target genes of Kit 1 but were ruled negative as per the Kit 1 criteria. Therefore, it can be inferred that using additional detection probes might be required to determine any false negative results obtained with a single kit. While such factors will always be a consideration in WBE approach and can only be resolved by upgrading the detection infrastructure, using more than one kit seems to increase the coverage of detection by 13% approximately which becomes more relevant as the increase in the number of samples becomes larger. It is worth highlighting that gene N, while could be detected by Kit 1 probe very efficiently, did have a couple of instances of only being detected by the probe from Kit 2 only indicating the benefits of multiple target approach.

It is interesting to note that not all the target genes could be detected uniformly or independently in all the samples analysed. This observation further highlights the need of analysis of multiple targets while surveilling pathogen presence in a given community. In addition to the use of multiple targets for detection, the criteria for considering a sample positive in terms of wastewater also needs to be revisited owing to the dilution of the virus particle and/or genes in the wastewater samples or the presence of various inhibitors. The current study for instance has followed the ICMR approved criteria of detecting two out of three targets for a sample to be positive by Kit 1. However, it is worth noting that these criteria were first approved for testing in patients directly and might not be as applicable to the wastewater sample where the presence of a single gene (with Ct values below the detection cut-off of 40) might indicate a very low presence of circulating pathogens in the community. Such observations should be worth a second look as even though it is possible that they are false positives, an alternative scenario where the detection of a spread is inhibited by some community specific factors which might be present in the wastewater samples. Standardizing a way therefore where the validity of presence of one single target out of many may provide a more sensitive application of WBE in containing a large-scale spread.

### 4.3. Quantification by two different targets probes could cover the complete second wave in the city

It was also interesting to note that even though the sensitivity of two quantitative probes seemed to be different (Limit of detection for N gene being log 10^3^ Genome concentration while that of ORF1ab being Log 10^4^) (Figure 4), both the genes could be quantified throughout the study period. Although a more sensitive detection method like droplet PCRs in combination to an integrated wastewater-based monitoring is ideal for the monitoring at the city level, this study has tried to investigate the success of WBE monitoring on the existing current wastewater treatment facilities in the city using simple qPCR-based detection. It was observed that with a weekly monitoring across the selected few sites in the city of Jaipur (9 sites spread across the city-7 longitudinally and 2 cross sectionally) the data of sites’ sample positivity could be correlated with a rise in active case rate approximately 14-20 days in advance. This observation has great implications in the context of countries like India where dense population per unit area and minimal individual testing facilities are real limitations. Indeed, the lack of applications like wastewater-based epidemiology predictions in city-wide pandemic management was painfully apparent during the second wave of COVID-19 in Jaipur. The study clearly hypothesizes that with the appropriate individual testing and smart lockdown strategy including micro-containment zone formation based on WBE prediction observations, unnecessary loss of many lives could have been saved along with reduced burden on the healthcare sector by proper management of the resources leading to reduced mortality & morbidity rate.

### 4.4. High SARS-CoV-2 loading led to incomplete removal in WWTPs

It is well known that these WWTPs are designed as per certain design parameters & criteria and work on the specified limited capacity. During the first wave of COVID-19 in Jaipur, it was observed that the treatment technologies available at different WWTPs in the city were sufficient in the removal of SARS-CoV-2 genome from the effluents and none of the effluent samples in the 14WWTPs were detected positive by qualitative assays (Arora et al 2021). However, during the second wave, it was observed that 37% (20/54) effluent samples were tested positive for the target genes both by qualitative and quantitative assays. This observation can be explained by several factors. Firstly, during the early phase of COVID-19 infections in 2020, the samples were collected during the months of May-July when the number of daily confirmed cases were in the range of 40-45 cases per day, while during the second wave in city, the daily confirmed case numbers were as high as 4202 (on May 82021) which is approximately 100 times higher. Thus, it can be extrapolated that the shear load of viral particles shed in the wastewater has increased significantly and possibly even higher than the working capacities of the treatment plants. This suggests a clear correlation between increased load in influent and RNA decay efficiency. The duration of the effluent sample collection corresponds to the months (April & May 2021) of maximum patient case load in the city. Secondly, it was also observed that upon exceeding a load of 5 log genome copies (GC) in the influent; quantification of gene load of neither gene N nor ORF1ab seemed to be proportional to their corresponding influent loads. This is an interesting observation in terms of RNA decay efficiency as well as retention. Thus, a) it is possible that during the peak phase of the second wave the wastewater treatment systems were exceedingly overloaded with the viral genome that they simply failed in complete removal of the viral RNA fragments; b) the uniform quantities of gene loads within their median load indicates that there is a steady retention of the genomic fragments in the treatment system reaching saturation under high viral load or c) the difference in two of the tertiary treated samples in case of the existence of the second mechanism it would be interesting to find out which component of treatment might promote this retention and is there any possibility that viral particles can be viable for a prolonged duration in these retended fractions. Further, it is important to understand the size of the treatment plant and operational and management consistencies, along with the quality of influent water will play a critical role in understanding in depth about the entire research scenario of COVID-19 transmission and monitoring.

Further, it is imperative to understand sample collection was done from three different secondary treatment technologies viz., ASP, MBBR and SBR and followed by two tertiary disinfection processes (UV and Chlorine) and still found the genetic fragments of SARS-CoV-2 in the effluent. This observation may imply that owing to nano-sized colloidal nature of genetic fragments, disinfection processes like chlorination/UV are likely to be less effective than the process of coagulation as reported in Kumar et al., 2021. In our study, all the different treatment processes are found to effectively remove SARS-CoV-2 RNA with varying efficacy. To the best of our knowledge, this is the first report assessing the effectiveness of five different treatment schemes for SARS-CoV-2 RNA reduction. As far as treatment type is concerned, the (SBR + Cl_2_) showed better efficacy of 81.2%, followed by MBBR + UV with 68.8% followed by SBR, MBBR + Cl_2_ and ASP. However, the detection & quantification of SARS-CoV-2RNAin wastewater does not imply viable viruses, it is highly recommended to validate on the infectivity/viability of SARS-CoV-2 after the treatment. However, it is worth considering here that effective aerobic WWTPs may not be sufficient to completely removes the genetic fragments of SARS-CoV-2.

### 4.5. Relevance of standardizing the WBE protocols& guidelines for pandemic management in a city like Jaipur

As mentioned above, the application of WBE in surveillance and monitoring of physiological and pathogenic trace markers has attained a lot of attention in countries with fully developed and integrated wastewater treatment systems during the COVID-19 pandemic. However, more research is required before this model can be adopted at a city-wide or national level, beyond the pandemic era particularly in developing countries. In India, these applications are even further from application owing to the limitations faced by underdeveloped wastewater treatment systems.

First, despite being a cost-effective measure to complement the individual testing; state of current sanitation network is thought of as a major challenge. In this context, establishing sophisticated sewerage systems will become an expensive step. This glaring gap, along with the consequent lack of awareness in policy makers of India has led to a nationwide reluctance in developing WBE methods in India. This study thus provides the example of successful application of WBE in Jaipur, a city with fragmented sewerage system. Even if it is possible to detect an upcoming wave as early as 14-20 days, various requirements as per the internationally established protocols previously reported by other groups e.g. size of sample collected, cold chain transportation, facilities of ultrafiltration setups, ultracentrifugation, sophisticated detection instruments like droplet PCR, etc. still need to be established.

The focus of this study was to apply the procedure developed in the lab scale to the city surveillance. Therefore, a feasibility of using these steps over a full wave of SARS-CoV-2 surge in the city was analysed. The advantages of the method used here include less time consuming, lesser number of steps and very less equipment requirements. For example, the prediction could be done successfully with randomly taking just 1 ml volume from 1 litre grab samples indicating that there is no need for large collection volume saving the transportation cost. Also, these samples were collected and transported to the laboratory at the ambient city temperatures (non-refrigerated vehicles, duration between collection and storage at 4 degrees was maximum 3.5 hours) for same day RNA isolation and qualitative detection suggesting cheaper sample collection and storage on a city level. Along with the observations of Arora et al., 2021, this collection model can be extended to remote locations where facilities of RNA extraction and detection might not be established. Therefore, instead of establishing and maintaining testing centres in every small village, gated community or towns, the collected sample can simply be transported to a centralized testing centre under cold chain-transport for detection and monitoring on a regular basis. The protocol for sample pre-processing is simple and can be completed by using even low speed centrifuges. Thus, the procedure reported in this study has been shown to be working perfectly on city scale weekly monitoring and can be applied to scale up even harnessing in the moderately equipped cities with centrally equipped detection centres.

## 5. Conclusions

Wastewater surveillance is a promising tool that detects real-time and early disease signals & determines emerging hotspots in the surveillance of COVID-19 prevalence at the community level. Yet in India, the city scale surveillance of SARS-CoV-2 RNA in wastewater remains poorly understood and needs to be explored, especially in cities with fragmented sewerage systems. A temporal variation of SARS-CoV-2 RNA presence in wastewater was studied for a period of five months in Jaipur, India. This study reported the first successful SARS-CoV-2 WBE application in 9 wastewater systems in Jaipur (n = 164) with varying sizes, which serve 60-70% of Jaipur’s sewerage network). Interestingly, the positive detection rates of SARS-CoV-2 RNA in the wastewater from all the WWTPs increased along with the clinical cases over time. A total of 72 samples (43.9%) of the total 164 samples tested in the study were found to be positive, with at least two positive RT-PCR results targeting four SARS-CoV-2 genes such as E, RdRp, N and ORF1ab gene. This system of wastewater-based epidemiology is extremely essential in practice in an Indian context where the resources are lacking in terms of both disease management and diagnosis. As demonstrated by this study, a gap of 14-20 days warning could be sufficient to take necessary actions to stop the spread of the next COVID-19 wave. This finding was further supported by the relation between the percentage change in effective gene concentration level and confirmed cases, which followed a similar trend on the temporal scale with a ∼1 to 2 weeks’ time distance. The study has successfully proven the global implications of WBE for India, highlighting the role of WBE through application of scalable and cost-effective protocol reported in the study for societal benefit and third wave improved management.

## Data Availability

All the necessary Data is available in the manuscript.

## Acknowledgements

The study group would like to acknowledge the constant support received from Dr. B. Lal Gupta (Director) and Dr. Aparna Datta (Principal). The support received from the Centre for Innovation, Research & development (CIRD, Dr. B. Lal Clinical laboratory Pvt. Ltd.) for analysis and the Jaipur development authority (JDA) officials & plant operators at WWTPs is sincerely acknowledged.

## Credit Author Statement

**Sudipti Arora:** Conceptualization, Methodology, Validation, Formal analysis, Investigation, Resources, Writing – Original Draft, Writing – Review & Editing, Project administration, Funding acquisition.

**Aditi Nag:** Conceptualization, Formal analysis, Investigation, Writing – Original Draft, Writing – Review & Editing, Visualization.

**Aakanksha Kalra:** Formal analysis, Visualization.

**Vikky Sinha**: Investigation.

**Ekta Meena**: Investigation.

**Samvida Saxena**: Investigation.

**Devanshi Sutaria**: Investigation, Writing – Review & Editing

**Manpreet Kaur**: Investigation.

**Tamanna Pamnani**: Investigation.

**Komal Sharma**: Investigation.

**Sonika Saxena**: Resources, Supervision, Project administration

**Sandeep K Shrivastava:** Resources, Methodology, Validation

**AB. Gupta**: Conceptualization, Formal analysis, Writing – Review & Editing

**Xuan Li**: Formal analysis, Writing – Review & Editing

**Guangming Jiang:** Formal analysis, Writing – Review & Editing

